# The incubation period of COVID-19: A scoping review and meta-analysis to aid modelling and planning

**DOI:** 10.1101/2020.10.20.20216143

**Authors:** Prakashini Banka, Catherine Comiskey

**Affiliations:** School of Nursing and Midwifery, Trinity College Dublin

**Keywords:** COVID-19, Self-isolation, Incubation Period, Asymptomatic Infections, Prevalence, Pre-Symptomatic

## Abstract

**Background:** An accurate estimate of the distribution of the incubation period for COVID-19 is the foundational building block for modelling the spread of the SARS COV2 and the effectiveness of mitigation strategies on affected communities. Initial estimates were based on early infections, the aim of this study was to provide an updated estimate and meta-analysis of the incubation period distribution for COVID-19.

**Methods:** The review was conducted according to the PRISMA Scoping Review guidelines. Five databases were searched; CINAHL, MEDLINE, PUBMED, EMBASE, ASSIA, and Global Index Medicus for studies published between 1 January 2020 - 27 July 2020.

**Results:** A total of 1,084 articles were identified through the database searches and 1 article was identified through the reference screening of retrieved articles. After screening 64 articles were included. The studies combined had a sample of 45,151 people. The mean of the incubation periods was 6.71 days with 95% CIs ranging from 1 to 12.4 days. The median was 6 days and IQR ranging from 1.8 to 16.3. The resulting parameters for a Gamma Distribution modelling the incubation period were Γ(*α, λ*) = Γ(2.810,0.419) with mean, *μ* = *α*/*λ*.

**Conclusion:** Governments are planning their strategies on a maximum incubation period of 14 days. While our results are limited to primarily Chinese research studies, the findings highlight the variability in the mean period and the potential for further incubation beyond 14 days. There is an ongoing need for detailed surveillance on the timing of self-isolation periods and related measures protecting communities as incubation periods may be longer.

## INTRODUCTION

To define the incubation period ECDC (2012) provides a simple overview of infectious disease epidemiology definitions at the different stages of infection according to time. The period between exposure and onset of clinical symptoms is called the ‘incubation period’. The host may become infectious (i.e. able to transmit the pathogen to other hosts) at any moment of the infection and this moment will vary per pathogen^1^

Early estimates of the incubation period of the emerging SARS COV2 virus were reported by Backer et al in Wuhan, China on the 27^th^ January 2020^2^. Authors reported that within the cases observed the incubation period was estimated to be 6.4 days. Following on from this early estimate authors Li et al in Wuhan reported a mean incubation period of 5.2 days^3^ and Lauer et al in China reported the median of the incubation period of 5.1 days^4^. Further individual studies in Europe by Bohmer et al have shown that the median period was 4 days with an Interquartile period of 2 days^5^. Within Saudi Arabia it has been shown that the incubation period can be 6 days^6^. Since these early studies, numerous further individual cases notifications and modelling studies have emerged estimating the period to range from 1 to 34 days^2-65^. However, to date no scoping review or meta-analysis has been conducted on the results of these individual studies with the primary objective of quantifying the mean and standard deviation of the incubation period for the purposes of modelling it’s distribution.

In the early stages of a new epidemic where no vaccine is available all persons are susceptible particularly some communities of older and vulnerable people. As the epidemic progresses and the number of infectious individuals increases the number of susceptible individuals will decrease. However, when an epidemic can produce both asymptomatic and symptomatic cases the identification of the numbers infected becomes more challenging. Mathematical modelling of the effective reproductive number R of an epidemic, the spread of infection, and the subsequent decisions on planning and mitigation is fully dependent on accurate and up to date estimates of the key modelling parameters. To aid this process and given the passage of time and improvements in surveillance and monitoring since the first cases, we conducted a scoping review and meta-analysis of the parameters of the global incubation period of COVID-19. Our ultimate aim was to provide an up to date estimate of the incubation period distribution to aid ongoing modelling estimates of the effective reproductive number R and subsequent estimates of the hidden prevalence of asymptomatic cases.

## METHODS

The study design was a scoping review with a meta-analysis. Daudt et al^66^in their work on enhancing the scoping review methodology based on the seminal work on scoping reviews by Askey and O’Malley (2005) retain the original definition of a scoping review and state that such reviews, *“map rapidly the key concepts underpinning a research area and the main sources and types of evidence available, and can be undertaken as standalone projects in their own right, especially where an area is complex or has not been reviewed comprehensively before”*^66^p.2. Within this review we are seeking to rapidly uncover, ‘the main sources and types of evidence available in an area that has not been reviewed comprehensively before’. In addition, the framework Askey and O’Malley (2005) developed expanded on this definition by identifying four main reasons for conducting a scoping study: (1) to examine the extent, range and nature of research activity; (2) to determine the value of undertaking a full systematic review; (3) to summarise and disseminate research findings; and (4) to identify research gaps in the existing literature. Within this review aims three and four are considered^66^. Levac et al (2010) highlight that scoping studies differ from systematic reviews because authors do not typically assess the quality of included studies^67^. Scoping studies also differ from narrative or literature reviews in that the scoping process can require analytical reinterpretation of the literature. Grant et al in their typology of reviews describe scoping reviews as providing a preliminary assessment of the potential size and scope of available research in the literature, with the aim of identifying the nature and extent of research evidence including ongoing research^68^. The completeness of the search in a scoping review is also determined by time and scope constraints. They share several characteristics of the systematic review in attempting to be systematic, transparent and replicable however their lack of a quality assessment and their rapid duration are recognised limitations.

### Search strategy and selection criteria

This review was conducted according to the PRISMA Scoping Review guidelines^69^. Five databases were searched; CINAHL, MEDLINE, PUBMED, EMBASE, ASSIA, and Global Index Medicus for studies published between 1 January 2020 and 27 July 2020. No language restrictions were applied, and non-English publications were translated. Search terms used included “coronavirus”, “covid-19”, AND “Incubation.” An example of search terms used for CINAHL was *(((MH “Coronavirus+”) or coronavirus*) AND (wuhan or beijing or shanghai or “2019-nCoV” or “Covid-19” or “SARS-CoV-2”)) OR ((((“novel coronavirus*” AND ((MH “China”) or China)) OR TI coronavirus*) OR (((MH pneumonia) or pneumonia) AND Wuhan OR ((“Covid-19” or “2019-nCoV” or “SARS-CoV-2” or (MH Coronavirus Infections))))) AND PY 2020) AND (MH Incubation)*.

The database search was supplemented by screening the references of retrieved articles. Data was imported into EndNote version x9 and Excel was also used for data extraction. Duplicates were removed using EndNote and another duplicate check was conducted manually. Studies were included if they presented incubation data on COVID-19. Study designs included retrospective case studies, multicentre studies, case series, prospective cohort studies, web data mining, cross-sectional studies, and individual case reports. Studies were excluded if they did not include findings on incubation period of COVID-19 or new information on the incubation period, or if they were reviews, no other study designs were excluded. Studies which assumed values of the incubation period based on other studies were also excluded at full text screening stage. Grey literature sources and unpublished studies were also assessed but trial registries were not.

One reviewer performed the search and the screening and queries were resolved with the second reviewer. Data were extracted based on the publication and no individual patient-level data were requested from the study authors, summary of estimates were included.

### Data analysis

Data extraction was conducted and summarised by PB. Duplicates were removed automatically using EndNote and a manual check was conducted. Extracted variables included first author, country, study design, sample size, and incubation period data with the primary outcomes being the mean, standard deviation and 95% confidence interval for the mean of the period and secondary outcomes being the minimum, maximum, range, median, interquartile range, and any additional reported confidence intervals. Using the Central Limit Theorem, the pooled mean was computed, and a pooled estimate of the variance and standard deviation were provided using standard formulas. For example, if we have 30 relevant studies we have,

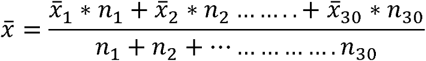

The standard deviation was available for four studies giving a pooled estimate described by,

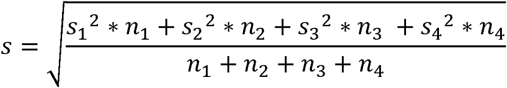

The median of the medians and the ranges in all other outcomes were provided. Forest plots were prepared to display the mean and median values and their respective 95% confidence intervals and inter-quartile ranges.

To model the distribution of incubation times in an epidemic a Gamma Probability Density Function is recommended^41^.This is described by

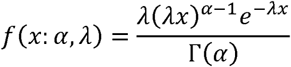

where *f* Is the probability of an incubation period of duration *x* given a mean incubation period of mean, *¼*= *±*/ *»*Parameter values for the Gamma Distribution describing the incubation period distribution were prepared based on the review findings.

As this was a scoping review a quality assessment of the studies was not carried out as is in keeping with the rapid nature and the protocol for this review type.

## RESULTS

A total of 1,084 articles were identified through the database searches and 1 article was identified through the reference screening of retrieved articles. Duplicates were removed (n= 227) and 858 articles were screened based on abstract and title. After the screening 689 articles were excluded and the remaining 169 articles were assessed for eligibility. The total number of full-text articles excluded with reasons was 105. A breakdown is provided in **Figure 1**.

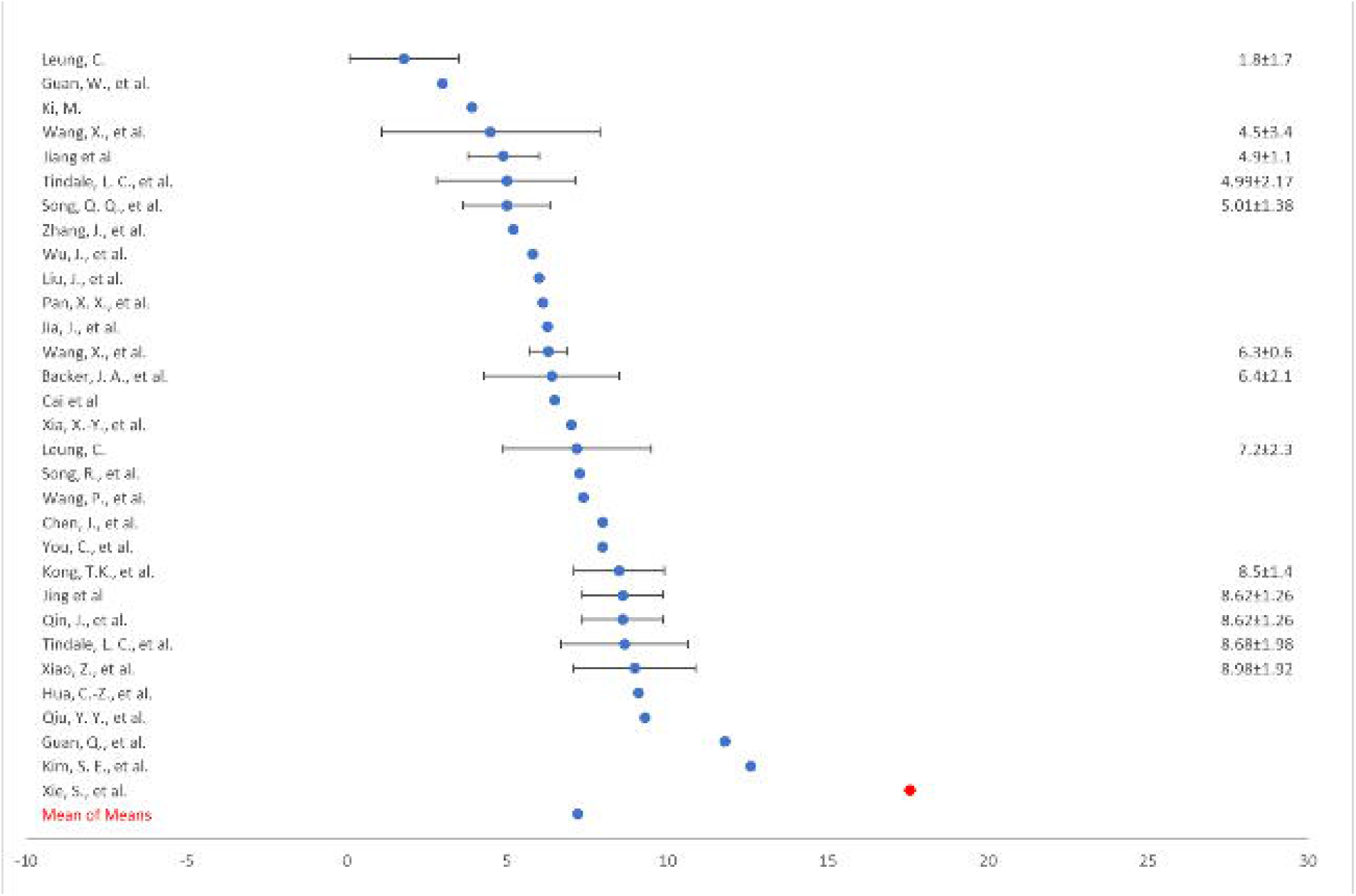

The 64 included studies had a combined sample size of 45,151 **(Table 1). Table 2** provides a summary of the incubation period from the 64 studies included in this scoping review. The age ranged between 0 and 90 years, with a median of 43 years and mean of 43.9 years. The minimum incubation period ranged between 1 and 9 days. The maximum incubation period ranged between 4 and 34 days. The mean incubation period was 6.71 (31 studies), and the median was 6 days (58 studies).

**Table 1:** Detailed Results from the 64 Articles

| Author | Country | n | Study Design | Groups | Min | Median | Max | Mean | SD | CI95% of the means |
| --- | --- | --- | --- | --- | --- | --- | --- | --- | --- | --- |
| Alsofayan, Y. M., et al. | Saudi Arabia | 1519 | Retrospective Cross-sectional | <56 |  | 6 |  |  |  |  |
|  |  |  |  | >56 |  | 13 |  |  |  |  |
| Backer, J. A., et al. | China | 88 | Retrospective Cross-sectional |  | 2.1 |  | 11.1 | 6.4 |  | 5.6 - 7.7 |
| Böhmer, M. M., et al. | Germany | 16 | Case Study Design |  | 1 | 4 | 7 |  |  |  |
| Chen, J., et al. | China | 12 | Retrospective Cross-sectional |  | 1 |  | 13 | 8 |  |  |
| Deng, Y., et al. | China |  | Retrospective Cross-sectional |  |  | 8.5 |  |  |  | 7.22 - 9.15 |
| Dong, X. C., et al. | China | 135 | Retrospective Cross-sectional |  | 3 | 6.5 | 13 |  |  |  |
| Gao, Y., et al. | China | 15 | Case Study Design |  | 3 | 10 | 12 |  |  |  |
| Gou, F. X., et al. | China | 63 | Retrospective Cross-sectional |  |  | 6 |  |  |  |  |
| Guan, Q., et al. | China | 8 | Case Study Design |  | 6 | 12.5 | 19 | 11.8 |  |  |
| Guan, W., et al. | China | 1099 | Retrospective Cross-sectional |  |  | 4 |  | 3 |  |  |
| Haiyan, Y., et al. | China | 1719 | Retrospective Cross-sectional |  | 1 | 7 | 20 |  |  |  |
| Han, Y.-N., et al. | China | 32 | Case Study Design | Children | 3 | 5 | 12 |  |  |  |
|  |  |  |  | Adult | 2 | 4 | 12 |  |  |  |
| Hua, C.-Z., et al. | China | 417 | Retrospective Multicentre Study | Children | 4 |  | 21 | 9.1 | 3.7 |  |
| Huang, L., et al. | China | 8 | Prospective Cohort Study |  | 1 | 2 | 4 |  |  |  |
| Jia, J., et al. | China | 44 | Retrospective Cross-sectional |  | 1 |  | 14 | 6.28 |  |  |
| Jin, X., et al. | China | 651 | Retrospective Cross-sectional | With GI Symptoms |  | 4 |  |  |  |  |
|  |  |  |  | Without GI Symptoms |  | 5 |  |  |  |  |
| Ki, M. | Korea | 28 | Retrospective Cross-sectional |  | 1 | 3 | 15 | 3.9 |  |  |
| Kim, S. E., et al. | South Korea | 13 | Case Study Design |  | 4 | 10 | 20 | 12.6 |  |  |
| Kong, D., et al. | China | 10 | Case Study Design |  | 1 | 6 | <14 |  |  |  |
| Kong, T. K. | China | 136 | Retrospective Cross-sectional | Combined | 1 | 8.3 | 17 | 8.5 |  | 7.8-9.2 |
|  |  |  |  | 15 - 64 | 1 | 7.6 | 17 |  |  |  |
|  |  |  |  | 65> | 3 | 11.2 | 17 |  |  |  |
| Lai, C. K. C., et al. | Hong Kong | 100 | Retrospective Cross-sectional |  |  | 4.2 |  |  |  | 4 - 2.5 |
| Lauer, S. A., et al. | China | 181 | Pooled Analysis |  |  | 5.1 |  |  |  | 4.5 - 5.8 |
| Lee, H., et al. | South Korea | 47 | Retrospective Cross-sectional |  |  | 3 |  |  |  |  |
| Leung, C. | China | 151 | Web Data Mining | Travellers (Weibull) |  |  |  | 1.8 |  | 1 - 2.7 |
|  |  |  |  | Non-Travellers (Weibull) |  |  |  | 7.2 |  | 6.1 - 8.4 |
| Li, J., et al. | China | 74 | Retrospective Cross-sectional |  |  | 5 |  |  |  |  |
| Li, Q., et al. | China | 425 | Retrospective Cross-sectional |  |  | 5.2 |  |  |  | 4.1 - 7 |
| Linton, N. M., et al. | China | 52 | Retrospective Cross-sectional |  | 2 | 5.6 | 14 |  |  | 4.4 - 7.4 |
| Liu, F., et al. | China | 10 | Retrospective Cross-sectional |  | 3 |  | 7 |  |  |  |
| Liu, J., et al. | China | 365 | Retrospective Cross-sectional |  | 1 | 5 | 16 | 6 |  |  |
| Nie, X., et al. | China | 7015 | Retrospective Cross-sectional |  | 2 | 5 | 15 |  |  |  |
| Pan, X. X., et al. | China | 52 | Retrospective Cross-sectional |  | 2 |  | 12 | 6.11 | 3.38 |  |
| Pung, R., et al. | Singapore | 17 | Retrospective Cross-sectional |  |  | 4 |  |  |  |  |
| Qian, G. Q., et al. | China | 91 | Retrospective Multicentre Study |  |  | 6 |  |  |  |  |
| Qin, J., et al. | China | 12963 | Retrospective Cross-sectional<br>and forward follow-up study |  |  | 8.13 |  | 8.62 |  | 8.02 - 9.28 |
| Qiu, C., et al. | China | 104 | Case Study Design |  | 1 | 6 | 32 |  |  |  |
| Qiu, Y. Y., et al. | China | 8 | Case Study Design |  | 8 | 9.5 | 11 | 9.3 |  |  |
| Shen, Q., et al. | China | 9 | Case Study Design |  | 1 | 7.5 | 16 |  |  |  |
| Shen, Y., et al. | China | 539 | Case-contact study |  | 4 | 7 | 12 |  |  |  |
| Song, Q. Q., et al. | China |  | Retrospective Cross-sectional |  |  |  |  | 5.01 |  | 4.31 - 5.69 |
| Song, R., et al. | China | 24 | Case Study Design |  | 2 | 8 | 13 | 7.27 |  |  |
| Sun, L., et al. | China | 55 | Retrospective Cross-sectional |  |  | 7.5 |  |  |  |  |
| Tian, S., et al. | China | 262 | Retrospective Cross-sectional |  |  | 6.7 |  |  |  |  |
| Tindale, L. C., et al. | Singapore | 93 | Retrospective Cross-sectional | Singapore |  | 5.32 |  | 4.99 |  | 4.97 - 7.14 |
|  | China | 135 |  | China |  | 8.06 |  | 8.68 |  | 7.72 - 9.7 |
| Wang, F., et al. | China | 333 | Retrospective Cross-sectional |  | 4 | 7 | 12 |  |  |  |
| Wang, P., et al. | China | 1212 | Retrospective Cross-sectional |  |  | 7 |  | 7.4 |  |  |
| Wang, X., et al. | China | 585 | Retrospective Cross-sectional |  |  | 5.7 |  | 6.3 |  | 6 - 6.6 |
| Wang, X., et al. | China | 35 | Retrospective Cross-sectional |  | 1 | 4 | 12 | 4.5 |  | 3 - 6.4 |
| Wang, Y., et al. | China | 275 | Retrospective Cross-sectional |  |  | 6 |  |  |  |  |
| Wu, J., et al. | China | 83 | Prospective Cohort Study |  |  | 4.3 |  | 5.8 |  |  |
| Wu, W. S., et al. | China | 40 | Retrospective Cross-sectional |  | 3 | 6 | 13 |  |  |  |
| Xia, X.-Y., et al. | China | 10 | Case Study Design |  | 2 |  | 14 | 7 | 2.59 |  |
| Xiao, Z., et al. | China | 2555 | Retrospective Cross-sectional |  |  | 8 |  | 8.98 |  | 7.98 - 9.9 |
| Xie, S., et al. | China | 105 | Retrospective Multicentre Study |  | 5 |  | 34 | 17.6 |  |  |
| Xu, T., et al. | China | 51 | Retrospective Cross-sectional | Imported | 4 | 8 | 11 |  |  |  |
|  |  |  |  | Secondary | 9 | 12 | 14 |  |  |  |
| Xu, X.-W., et al. | China | 62 | Case Study Design |  |  | 4 |  |  |  |  |
| Yang, H. Y., et al. | China | 325 | Case Study Design |  | 1 | 7 | 20 |  |  |  |
| Yang, L., et al. | China | 178 | Retrospective Case Series |  |  | 5.4 |  |  |  |  |
| You, C., et al. | China | 198 | Retrospective Cross-sectional |  |  | 7 |  | 8 | 4.75 |  |
| Yu, X., et al. | China | 333 | Retrospective Cross-sectional |  |  | 7.2 |  |  |  |  |
| Zhang, J., et al. | China | 8579 | Retrospective Cross-sectional |  |  |  |  | 5.2 |  | 1.8 - 12.4 |
| Zhao, C., et al. | China | 136 | Retrospective Case Series |  |  | 6 |  |  |  |  |
| Jiang et al | China | 50 | Retrospective Cross-sectional |  |  |  |  | 4.9 |  | 4.4 - 5.5 |
| Cai et al | China | 10 | Case Study Design |  | 2 |  | 10 | 6.5 |  |  |
| Jing et al | China | 1211 | Retrospective Cross-sectional |  |  | 8.13 |  | 8.62 |  | 8.02 - 9.28 |

**Table 2:** Summary of Incubation Period

| N= 45,151 | Range | Median | Pooled Estimate |
| --- | --- | --- | --- |
| Age | 0.00 - 90.0 | 43.0 | - |
| Minimum | 1.0 - 9.0 | - | - |
| Maximum | 4.0 - 34.0 | - | - |
| Median | 2.0 - 13.0 | 6.0 | - |
| Interquartile Range of the Medians | 1.8 – 16.3 | - | - |
| Mean | 1.8 – 17.6 | - | 6.71 |
| Standard Deviation | 2.59 - 4.75 | - | 4.00 |
| Confidence Interval of the Means | 1.0 - 12.4 | - | - |
| Mean and variance of the Gamma distribution $\bar{x} = \frac{\alpha}{\lambda}$ and $s^2 = \frac{\alpha}{\lambda^2}$ | | | |

Forest plot illustrating the spread in the mean values, and their respective confidence intervals are provided in Figures 2. We can see from Figure 2 that 30 studies provided information on the mean value of the observed incubation period. These thirty studies represented a combined sample size of 43,132 individuals. Only 12 of the 30 studies provided estimates for the 95% confidence interval of the mean incubation period. A total of 58 studies provided details on the median incubation period observed and 18 of these provided some details on the interquartile range. Given the pooled estimates for the mean and the standard deviation and given the Gamma Distribution where 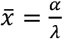 and 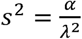 describes the incubation period distribution we found that the shape parameter was given *α*=2.81 and *λ*=0.42 was the estimated rate or scale parameter.

## DISCUSSION

This scoping review finds that based on studies to date, the mean incubation period for COVID-19 is given by 6.71 days with a standard deviation of 4.00 days and 95% CIs ranging from 1.8 to 12.4 days. These findings provide for the first time the parameter estimates required for a Gamma distribution to model the period.

The importance of accurately modelling the incubation period on the spread of an epidemic has long been identified from the earliest work of Italian physician Girolamo Fracastro (1478-1553) on rabies ^70^ to the problem of modelling the incubation period of infectious diseases such as measles, poliomyelitis, smallpox and SARS ^70 71^. The importance of accurately measuring and modelling this period for SARS COV2 was identified early in this pandemic by Backer et al ^2^ when the mean incubation period was estimated to be 6.4 days and by Li et al ^3^ where a mean incubation period of 5.2 days was reported. Our updated review finds the mean may be longer than previously estimated and in addition provides the first estimates for a gamma distribution describing the period.

A limitation of our results is however the lack of published data on the variability in mean times. Of the 64 papers included in the review only 30 reported mean incubation period times and only 4 reported standard deviations or variance. This lack of information on the variability in mean times will undoubtedly introduce a level of uncertainty and bias into the Gamma distribution parameter estimates. Furthermore, the range in mean values reported across the 30 studies varied greatly. Clearly the quality of some studies was lacking, and this assessment of quality was by definition outside the stated objectives of a scoping g review. None the less, this scoping review has provided the first meta-analysis of 64 studies published from January to July 2020.

In spite of the provision of 95% confidence intervals for the mean incubation period it was not clear from the studies how these intervals were computed and estimates of the variance and standard deviation could not be computed from them using standard formulae. Given the lack of clarity on the variance and hence standard deviation it is recommended that in future research greater attention is given to the computation details and formula used are provided.

To conclude, for the planning and provision of mitigation strategies and advice on isolation periods for the general public, it is essential that an accurate and update estimate of the incubation period is maintained as the SARS COV2 virus spreads. It is equally essential that the mitigation strategies on affected communities and hospital planning requirements that are determined from modelling scenarios are based on reliable and accurate global estimates of the incubation period. There is an ongoing need for detailed surveillance on the timing of self-isolation periods and related measures protecting communities as incubation periods may be longer. This international scoping review provides this estimate based on the findings to the end of July 2020.

### What is already known on this subject

▸ Early estimates of the incubation period of the emerging SARS COV2 virus were reported by Backer et al in Wuhan, China on the 27^th^ January 2020.
▸ Following on from this early estimate authors Li et al in Wuhan reported a mean incubation period of 5.2 days and Lauer et al in China reported the median of the incubation period of 5.1 days.
▸ Since these early studies, numerous further individual cases notifications and modelling studies have emerged estimating the period to range from 1 to 34 days.

### What this study adds

▸ This scoping review finds that based on studies to date, the mean incubation period for COVID-19 is given by 6.71 days with a standard deviation of 4.00 days and 95% CIs ranging from 1.8 to 12.4 days.
▸ These findings provide for the first time the parameter estimates required for a Gamma distribution to model the period.

## Data Availability

All relevant data is provided in the paper.

## Notes

### Competing Interest Statement

The authors have declared no competing interest.

### Funding Statement

This research was funded by the Health Research Board and Irish Research Council under grant scheme COVID-19 Pandemic Rapid Response and grant number COV19-2020-010.

## REFERENCES

1. Danielsson N, Catchpole M. Novel coronavirus associated with severe respiratory disease: case definition and public health measures. Eurosurveillance 2012;17(39):20282.

2. Backer JA, Klinkenberg D, Wallinga J. Incubation period of 2019 novel coronavirus (2019-nCoV) infections among travellers from Wuhan, China, 20-28 January 2020. Euro surveillance : bulletin Europeen sur les maladies transmissibles = European communicable disease bulletin 2020;25(5) doi: 10.2807/1560-7917.ES.2020.25.5.2000062

3. Li Q, Guan X, Wu P, et al. Early Transmission Dynamics in Wuhan, China, of Novel Coronavirus-Infected Pneumonia. The New England journal of medicine 2020;382(13):1199–207. doi: 10.1056/NEJMoa2001316

4. Lauer SA, Grantz KH, Bi Q, et al. The Incubation Period of Coronavirus Disease 2019 (COVID-19) From Publicly Reported Confirmed Cases: Estimation and Application. Annals of Internal Medicine 2020;172(9):577–82. doi: 10.7326/M20-0504

5. Böhmer MM, Buchholz U, Corman VM, et al. Investigation of a COVID-19 outbreak in Germany resulting from a single travel-associated primary case: a case series. The Lancet Infectious diseases 2020 doi: 10.1016/S1473-3099(20)30314-5

6. Alsofayan YM, Althunayyan SM, Khan AA, et al. Clinical characteristics of COVID-19 in Saudi Arabia: A national retrospective study. Journal of Infection and Public Health 2020 doi: 10.1016/j.jiph.2020.05.026

7. Chen J, Zhang Z-Z, Chen Y-K, et al. The clinical and immunological features of pediatric COVID-19 patients in China. Genes & diseases 2020 doi: 10.1016/j.gendis.2020.03.008

8. Deng Y, You C, Liu Y, et al. Estimation of incubation period and generation time based on observed length-biased epidemic cohort with censoring for COVID-19 outbreak in China. Biometrics 2020 doi: 10.1111/biom.13325

9. Dong XC, Li JM, Bai JY, et al. Epidemiological characteristics of confirmed COVID-19 cases in Tianjin. Zhonghua liu xing bing xue za zhi = Zhonghua liuxingbingxue zazhi 2020;41(5):638–42. doi: 10.3760/cma.j.cn112338-20200221-00146

10. Gao Y, Shi C, Chen Y, et al. A cluster of the Corona Virus Disease 2019 caused by incubation period transmission in Wuxi, China. Journal of Infection 2020;80(6):666–70. doi: 10.1016/j.jinf.2020.03.042

11. Gou FX, Zhang XS, Yao JX, et al. Epidemiological characteristics of COVID-19 in Gansu province. Zhonghua liu xing bing xue za zhi = Zhonghua liuxingbingxue zazhi 2020;41:E032. doi: 10.3760/cma.j.cn112338-20200229-00216

12. Guan Q, Liu M, Zhuang YJ, et al. Epidemiological investigation of a family clustering of COVID-19. Zhonghua liu xing bing xue za zhi = Zhonghua liuxingbingxue zazhi 2020;41(5):629–33. doi: 10.3760/cma.j.cn112338-20200223-00152

13. Guan W, Ni Z, Hu Y, et al. Clinical characteristics of coronavirus disease 2019 in China. New England Journal of Medicine 2020;382(18):1708–20. doi: 10.1056/NEJMoa2002032

14. Haiyan Y, Jie XU, Yan LI, et al. The preliminary analysis on the characteristics of the cluster for the Corona Virus Disease. Chinese Journal of Epidemiology 2020(12):623–28.

15. Han Y-N, Feng Z-W, Sun L-N, et al. A comparative-descriptive analysis of clinical characteristics in 2019-coronavirus-infected children and adults. Journal of medical virology 2020 doi: 10.1002/jmv.25835

16. Hua C-Z, Miao Z-P, Zheng J-S, et al. Epidemiological features and viral shedding in children with SARS-CoV-2 infection. Journal of medical virology 2020 doi: 10.1002/jmv.26180

17. Huang L, Zhang X, Zhang X, et al. Rapid asymptomatic transmission of COVID-19 during the incubation period demonstrating strong infectivity in a cluster of youngsters aged 16-23 years outside Wuhan and characteristics of young patients with COVID-19: A prospective contact-tracing study. Journal of Infection 2020;80(6):e1–e13. doi: 10.1016/j.jinf.2020.03.006

18. Jia J, Hu X, Yang F, et al. Epidemiological characteristics on the clustering nature of COVID-19 in Qingdao City, 2020: a descriptive analysis. Disaster medicine and public health preparedness 2020:1–17. doi: 10.1017/dmp.2020.59

19. Jin X, Lian J-S, Hu J-H, et al. Epidemiological, clinical and virological characteristics of 74 cases of coronavirus-infected disease 2019 (COVID-19) with gastrointestinal symptoms. Gut 2020;69(6):1002–09. doi: 10.1136/gutjnl-2020-320926

20. Ki M. Epidemiologic characteristics of early cases with 2019 novel coronavirus (2019-nCoV) disease in Korea. Epidemiology & Health 2020;42:e2020007–e07. doi: 10.4178/epih.e2020007

21. Kim SE, Jeong HS, Yu Y, et al. Viral kinetics of SARS-CoV-2 in asymptomatic carriers and presymptomatic patients. International journal of infectious diseases : IJID : official publication of the International Society for Infectious Diseases 2020;95:441–43. doi: 10.1016/j.ijid.2020.04.083

22. Kong D, Zheng Y, Wu H, et al. Pre-symptomatic transmission of novel coronavirus in community settings. Influenza and other respiratory viruses 2020 doi: 10.1111/irv.12773

23. Kong T-K. Longer incubation period of coronavirus disease 2019 (COVID-19) in older adults. Aging medicine (Milton (NSW)) 2020;3(2):102–09. doi: 10.1002/agm2.12114

24. Lai CKC, Ng RWY, Wong MCS, et al. Epidemiological characteristics of the first 100 cases of coronavirus disease 2019 (COVID-19) in Hong Kong Special Administrative Region, China, a city with a stringent containment policy. International journal of epidemiology 2020 doi: 10.1093/ije/dyaa106

25. Lee H, Kim K, Choi K, et al. Incubation period of the coronavirus disease 2019 (COVID-19) in Busan, South Korea. Journal of infection and chemotherapy : official journal of the Japan Society of Chemotherapy 2020 doi: 10.1016/j.jiac.2020.06.018

26. Leung C. The difference in the incubation period of 2019 novel coronavirus (SARS-CoV-2) infection between travelers to Hubei and nontravelers: The need for a longer quarantine period. Infection Control & Hospital Epidemiology 2020;41(5):594–96. doi: 10.1017/ice.2020.81

27. Li J, Ding J, Chen L, et al. Epidemiological and clinical characteristics of three family clusters of COVID-19 transmitted by latent patients in China. Epidemiology and infection 2020;148:e137. doi: 10.1017/S0950268820001491

28. Linton NM, Kobayashi T, Yang Y, et al. Incubation Period and Other Epidemiological Characteristics of 2019 Novel Coronavirus Infections with Right Truncation: A Statistical Analysis of Publicly Available Case Data. Journal of clinical medicine 2020;9(2) doi: 10.3390/jcm9020538

29. Liu F, Xu A, Zhang Y, et al. Patients of COVID-19 may benefit from sustained Lopinavir-combined regimen and the increase of Eosinophil may predict the outcome of COVID-19 progression. International journal of infectious diseases : IJID : official publication of the International Society for Infectious Diseases 2020;95:183–91. doi: 10.1016/j.ijid.2020.03.013

30. Liu J, Liao X, Qian S, et al. Community Transmission of Severe Acute Respiratory Syndrome Coronavirus 2, Shenzhen, China, 2020. Emerging infectious diseases 2020;26(6):1320–23. doi: 10.3201/eid2606.200239

31. Nie X, Fan L, Mu G, et al. Epidemiological Characteristics and Incubation Period of 7015 Confirmed Cases With Coronavirus Disease 2019 Outside Hubei Province in China. Journal of Infectious Diseases 2020;221:26–33. doi: 10.1093/infdis/jiaa211

32. Pan XX, Chen Y, Wang AH, et al. Study on transmission dynamic of 15 clusters of coronavirus disease 2019 cases in Ningbo. Zhonghua liu xing bing xue za zhi = Zhonghua liuxingbingxue zazhi 2020;41:E066. doi: 10.3760/cma.j.cn112338-20200330-00466

33. Pung R, Chiew CJ, Young BE, et al. Investigation of three clusters of COVID-19 in Singapore: implications for surveillance and response measures. The Lancet 2020;395(10229):1039–46. doi: 10.1016/S0140-6736(20)30528-6

34. Qian GQ, Yang NB, Ding F, et al. Epidemiologic and clinical characteristics of 91 hospitalized patients with COVID-19 in Zhejiang, China: a retrospective, multi-centre case series. QJM : monthly journal of the Association of Physicians 2020;113(7):474–81. doi: 10.1093/qjmed/hcaa089

35. Cai J, Sun W, Huang J, et al. Indirect Virus Transmission in Cluster of COVID-19 Cases, Wenzhou, China, 2020. Emerging infectious diseases 2020;26(6):1343–45. doi: 10.3201/eid2606.200412

36. Qin J, You C, Lin Q, et al. Estimation of incubation period distribution of COVID-19 using disease onset forward time: a novel cross-sectional and forward follow-up study. MedRxiv : the preprint server for health sciences 2020 doi: 10.1101/2020.03.06.20032417

37. Qiu C, Deng Z, Xiao Q, et al. Transmission and clinical characteristics of coronavirus disease 2019 in 104 outside-Wuhan patients, China. Journal of medical virology 2020 doi: 10.1002/jmv.25975

38. Qiu YY, Wang SQ, Wang XL, et al. Epidemiological analysis on a family cluster of COVID-19. Zhonghua liu xing bing xue za zhi = Zhonghua liuxingbingxue zazhi 2020;41(4):494–97. doi: 10.3760/cma.j.cn112338-20200221-00147

39. Shen Q, Guo W, Guo T, et al. Novel coronavirus infection in children outside of Wuhan, China. Pediatric pulmonology 2020;55(6):1424–29. doi: 10.1002/ppul.24762

40. Shen Y, Xu W, Li C, et al. A Cluster of Novel Coronavirus Disease 2019 Infections Indicating Person-to-Person Transmission Among Casual Contacts From Social Gatherings: An Outbreak Case-Contact Investigation. Open forum infectious diseases 2020;7(6):ofaa231. doi: 10.1093/ofid/ofaa231

41. Song QQ, Zhao H, Fang LQ, et al. Study on assessing early epidemiological parameters of COVID-19 epidemic in China. Zhonghua liu xing bing xue za zhi = Zhonghua liuxingbingxue zazhi 2020;41(4):461–65. doi: 10.3760/cma.j.cn112338-20200205-00069

42. Song R, Han B, Song M, et al. Clinical and epidemiological features of COVID-19 family clusters in Beijing, China. The Journal of infection 2020 doi: 10.1016/j.jinf.2020.04.018

43. Sun L, Shen L, Fan J, et al. Clinical features of patients with coronavirus disease 2019 from a designated hospital in Beijing, China. Journal of medical virology 2020 doi: 10.1002/jmv.25966

44. Tian S, Hu N, Lou J, et al. Characteristics of COVID-19 infection in Beijing. Journal of Infection 2020;80(4):401–06. doi: 10.1016/j.jinf.2020.02.018

45. Tindale LC, Stockdale JE, Coombe M, et al. Evidence for transmission of covid-19 prior to symptom onset. eLife 2020;9:1–34. doi: 10.7554/eLife.57149

46. Wang F, Qu M, Zhou X, et al. The timeline and risk factors of clinical progression of COVID-19 in Shenzhen, China. Journal of Translational Medicine 2020;18(1) doi: 10.1186/s12967-020-02423-8

47. Wang P, Lu J-A, Jin Y, et al. Statistical and network analysis of 1212 COVID-19 patients in Henan, China. International journal of infectious diseases : IJID : official publication of the International Society for Infectious Diseases 2020;95:391–98. doi: 10.1016/j.ijid.2020.04.051

48. Wang X, Pan Y, Zhang D, et al. Basic epidemiological parameter values from data of real-world in mega-cities: the characteristics of COVID-19 in Beijing, China. BMC Infectious Diseases 2020;20(1):1–11. doi: 10.1186/s12879-020-05251-9

49. Wang X, Zhou Q, He Y, et al. Nosocomial outbreak of COVID-19 pneumonia in Wuhan, China. The European respiratory journal 2020;55(6) doi: 10.1183/13993003.00544-2020

50. Wang Y, Liao B, Guo Y, et al. Clinical Characteristics of Patients Infected With the Novel 2019 Coronavirus (SARS-Cov-2) in Guangzhou, China. Open forum infectious diseases 2020;7(6):ofaa187. doi: 10.1093/ofid/ofaa187

51. Wu J, Huang Y, Tu C, et al. Household Transmission of SARS-CoV-2, Zhuhai, China, 2020. Clinical infectious diseases : an official publication of the Infectious Diseases Society of America 2020 doi: 10.1093/cid/ciaa557

52. Wu WS, Li YG, Wei ZF, et al. Investigation and analysis on characteristics of a cluster of COVID-19 associated with exposure in a department store in Tianjin. Zhonghua liu xing bing xue za zhi = Zhonghua liuxingbingxue zazhi 2020;41(4):489–93. doi: 10.3760/cma.j.cn112338-20200221-00139

53. Xia X-Y, Wu J, Liu H-L, et al. Epidemiological and initial clinical characteristics of patients with family aggregation of COVID-19. Journal of clinical virology : the official publication of the Pan American Society for Clinical Virology 2020;127:104360. doi: 10.1016/j.jcv.2020.104360

54. Xiao Z, Xie X, Guo W, et al. Examining the incubation period distributions of COVID-19 on Chinese patients with different travel histories. Journal of infection in developing countries 2020;14(4):323–27. doi: 10.3855/jidc.12718

55. Xie S, Zhang G, Yu H, et al. The epidemiologic and clinical features of suspected and confirmed cases of imported 2019 novel coronavirus pneumonia in north Shanghai, China. Annals of translational medicine 2020;8(10):637. doi: 10.21037/atm-20-2119

56. Xu T, Chen C, Zhu Z, et al. Clinical features and dynamics of viral load in imported and non-imported patients with COVID-19. International journal of infectious diseases : IJID : official publication of the International Society for Infectious Diseases 2020;94:68–71. doi: 10.1016/j.ijid.2020.03.022

57. Xu X-W, Wu X-X, Jiang X-G, et al. Clinical findings in a group of patients infected with the 2019 novel coronavirus (SARS-Cov-2) outside of Wuhan, China: retrospective case series. BMJ (Clinical research ed) 2020;368:m606. doi: 10.1136/bmj.m606

58. Yang HY, Xu J, Li Y, et al. The preliminary analysis on the characteristics of the cluster for the Corona Virus Disease. Zhonghua liu xing bing xue za zhi = Zhonghua liuxingbingxue zazhi 2020;41:623–28. doi: 10.3760/cma.j.cn112338-20200223-00153

59. Yang L, Dai J, Zhao J, et al. Estimation of incubation period and serial interval of COVID-19: analysis of 178 cases and 131 transmission chains in Hubei province, China. Epidemiology and infection 2020;148:e117. doi: 10.1017/S0950268820001338

60. You C, Deng Y, Hu W, et al. Estimation of the time-varying reproduction number of COVID-19 outbreak in China. International Journal of Hygiene & Environmental Health 2020;228:N.PAG-N.PAG. doi: 10.1016/j.ijheh.2020.113555

61. Yu X, Sun X, Cui P, et al. Epidemiological and clinical characteristics of 333 confirmed cases with coronavirus disease 2019 in Shanghai, China. Transboundary and emerging diseases 2020;67(4):1697–707. doi: 10.1111/tbed.13604

62. Zhang J, Litvinova M, Wang W, et al. Evolving epidemiology and transmission dynamics of coronavirus disease 2019 outside Hubei province, China: a descriptive and modelling study. Lancet Infectious Diseases 2020;20(7):793–802. doi: 10.1016/S1473-3099(20)30230-9

63. Zhao C, Xu Y, Zhang X, et al. Public health initiatives from hospitalized patients with COVID-19, China. Journal of infection and public health 2020 doi: 10.1016/j.jiph.2020.06.013

64. Jiang X, Rayner S, Luo M-H. Does SARS-CoV-2 has a longer incubation period than SARS and MERS? Journal of medical virology 2020;92(5):476–78. doi: 10.1002/jmv.25708

65. Jing QL, Li YG, Ma MM, et al. Contagiousness and secondary attack rate of 2019 novel coronavirus based on cluster epidemics of COVID-19 in Guangzhou. Zhonghua liu xing bing xue za zhi = Zhonghua liuxingbingxue zazhi 2020;41:E058. doi: 10.3760/cma.j.cn112338-20200310-00305

66. Daudt HM, van Mossel C, Scott SJ. Enhancing the scoping study methodology: a large, inter-professional team’s experience with Arksey and O’Malley’s framework. BMC medical research methodology 2013;13(1):48.

67. Levac D, Colquhoun H, O’Brien KK. Scoping studies: advancing the methodology. Implementation science 2010;5(1):69.

68. Grant MJ, Booth A. A typology of reviews: an analysis of 14 review types and associated methodologies. Health Information & Libraries Journal 2009;26(2):91–108.

69. Tricco AC, Lillie E, Zarin W, et al. PRISMA extension for scoping reviews (PRISMA-ScR): checklist and explanation. Annals of internal medicine 2018;169(7):467–73.

70. Nishiura H. Early efforts in modeling the incubation period of infectious diseases with an acute course of illness. Emerging themes in epidemiology 2007;4(1):2.

71. Farewell VT, Herzberg A, James K, et al. SARS incubation and quarantine times: when is an exposed individual known to be disease free? Statistics in medicine 2005;24(22):3431–45.

